# Inhomogeneous mixing and asynchronic transmission between local outbreaks account for the spread of COVID-19 epidemics

**DOI:** 10.1101/2020.08.04.20168443

**Authors:** Carlos I. Mendoza

## Abstract

The ongoing epidemic of COVID-19 originated in China has reinforced the need to develop epidemiological models capable of describing the progression of the disease to be of use in the formulation of mitigation policies. Here, this problem is addressed using a metapopulation approach to show that the delay in the transmission of the spread between different subsets of the total population, can be incorporated into a SIR framework through a time-dependent transmission rate. Thus, the reproduction number decreases with time despite the population dynamics remains uniform and the depletion of susceptible individuals is small. The obtained results are consistent with the early subexponential growth observed in the cumulated number of confirmed cases even in the absence of containment measures. We validate our model by describing the evolution of the COVID-19 using real data from different countries with an emphasis in the case of Mexico and show that it describes correctly also the long-time dynamics of the spread. The proposed model yet simple is successful at describing the onset and progression of the outbreak and considerably improves accuracy of predictions over traditional compartmental models. The insights given here may probe be useful to forecast the extent of the public health risks of epidemics and thus improving public policy-making aimed at reducing such risks.

## Introduction

First detected in December 2019 in the city of Wuhan in Hubei province, China, the COVID-19 outbreak, caused by the newly identified coronavirus SARS-CoV-2 has spread around the globe and reached the status of pandemic on March 11th, 2020. Due to the severity of the damages to health it may cause and its ease of transmission, a number of different strategies have been implemented by the authorities of different countries to block or reduce the spread of the virus. In some cases like China, Italy or Spain, strict quarantine measures have been adopted [1], [2]. However, strict lockdown is in many cases impossible due to prevailing economical and social factors. In such cases, the authorities aimed for less strict mitigation policies [3] including social distancing and individual non-pharmaceutical interventions [4]. Nonetheless, an accurate description of the progression of an epidemics is of fundamental importance in helping to decide public policies to reduce its impact, specially to stay below a fixed healthcare capacity and delaying the peak of the epidemic so that healthcare capacity can be expanded to support patients.

In Mexico the first detected case of COVID-19 was registered on February 27th, 2020 and corresponded to an imported case from Italy. This marks the start of what was called phase 1 of the epidemics, characterized by imported cases only, with no local contagion. Mexican health authorities identified a total of three epidemiological phases, according to the degree of transmission of the disease. Phase 2 of the coronavirus pandemic, was characterized by cases of local contagion between people who have not had contact with foreigners. It was declared on March 24th and actions comprised primarily the suspension of certain economic activities, the suspension of lessons at schools, the restriction of mass congregations and the recommendation of domiciliary protection for the general population. As a consequence of the evolution of confirmed cases and deaths from the disease in the country, on March 30th, a "health emergency due to force majeure" was declared, which led to the execution of additional actions for its prevention and control, the most conspicuous was the generalized voluntary quarantine of the population (the so called, “Jornada Nacional de Sana Distancia”). Eventually, on April 21th, the phase 3, characterized by thousands of cases disseminated in all the country, was declared.

Intervention measures adopted with the intention to mitigate the spread, are normally based in estimates of the progression of the outbreak. Mathematical models of infectious diseases are important tools for assessing the threat of a novel pathogen and offer the best information for mitigating an outbreak [5], [6]. Hence the need for epidemiological models that are able to estimate with some degree of accuracy the evolution of the outbreak to help to evaluate the impact of interventions [7], [8], [9]. The paradigmatical approach traditionally used to model the dynamics of an epidemics is the well known SIR (Susceptible, Infectious, Removed) compartmental model [10], [11]. In this model, the group *S* represents individuals who are susceptible to the disease and can become infected, the group *I* represents individuals that are infectious and can infect susceptible individuals, and the group *R* represents removed individuals that either, gained life-long immunity at recovery or die; in either case, removed individuals can not infect or be infected anymore. Although successfully applied to describe the spread of an infectious agent in a *well-mixed* population [12], this same simplifying assumption prevents its successful application in many other cases [13] such as the recent outbreak of COVID-19 which shows early subexponential growth [14], [15]. In a well-mixed population, it is assumed an homogeneous distribution of the susceptible-infectious contacts such that any susceptible individual may be infected by any infectious individual in the whole population. However, there are many heterogeneities in human populations that influence virus transmission [16], thus, any realistic epidemics model should take them into account.

Given that the subexponential growth seems to be a generic characteristic of the COVID-19 outbreak, independently of the suppression strategies implemented to mitigate the temporal evolution of the epidemic process, it is suggested the existence of an underlying mechanism responsible for this temporal behavior. The purpose of the present study is to show that the asynchronicity of the epidemics of component subpopulations may be at the source of this behavior. It is also shown that the standard SIR model can be extended to include the above mentioned asynchronicity and that the resulting model captures correctly not only the short-time but also the long-time dynamics of the COVID-19 outbreaks.

## Incorporating asynchronic transmission in a SIR framework

A natural way to incorporate population heterogeneities or spatial structure into an infectious disease model is by means of metapopulation models [5], [17], [18]. Let us assume a metapopulation approach [18] in which the total population is considered as if it were formed by *n* identical subpopulations or patches connected to each other with transmission lines due to movements of individuals. In each subpopulation, an infectious agent spread is described by a standard SIR model with coupling terms

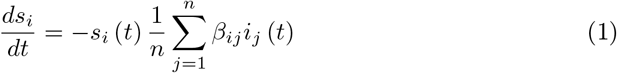

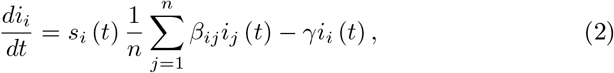

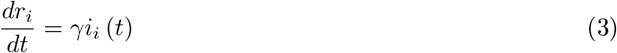

where *s_i_*(*t*) = *S_i_*(*t*)*/N_i_, i_i_*(*t*) *= I_i_*(*t*)*/N_i_*, and *r_i_*(*t*) *= R_i_*(*t*)*/N_i_* are the fractional susceptible, infectious and removed individuals of subpopulation *i* that has *N_i_* individuals. Since the population of each patch is assumed to be constant, then the fractional quantities satisfy the relationship *s_i_*(*t*) *+ i_i_*(*t*) *+ r_i_*(*t*) *=* 1. In Eqs. (1)-(3), *β_ij_* represents the elements of a matrix that describes the transmission between and within patches, the recovery rate γ is common to all subpopulations and, as in traditional SIR, all these quantities are time independent. The transmission rates *β_ij_* captures the rate of flow from group *S_i_* to group *I_j_* while the recovering rate γ indicates that infectious individuals get recovered of die at a fixed average rate γ. If subpopulations are independent from each other, then *β_ij_ = β*_0_δ*_ij_*, with *δ_ij_* the Kronecker delta function. The basic reproduction number *R*_0_ *= β*_0_/γ, captures the average number of secondary infections an infected will cause before he or she recovers or is effectively removed from the population. However, as the number of susceptible individuals in the population declines due to a growing number of infections, the effective reproduction number over time, *R_i,t_*, is given by the product of *R*_0_ and the fraction of susceptible individuals in the subpopulation *i*, *R_i,t_ = R*_0_*s_i_*(*t*). During the first few generations of disease transmission, in the absence of control interventions or reactive population behavior changes, the SIR model support a reproduction number that is essentially invariant, e.g., *R_i,t_ ≃ R*_0_. Thus, in the classical SIR model with constant transmission rate *β*_0_, in a completely susceptible population, *s_i_*(0) *≃* 1, *i_i_*(*t*) grows exponentially during the early epidemic phase [19], e.g., 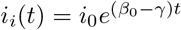.

It is unusual for a naturally occurring disease emergence to occur simultaneously at many locations. This means that at least during the initial phase of transmission, infectious individuals are clustered [7]. The presence of the subpopulations can be thought as clustered regions in space where a given individual spent most of his time. Clusters exchange pathogens with each other through infected or susceptible individuals traveling between them during the period of infectiousness. Thus, spatial inhomogeneities lead naturally to outbreaks that do not occur simultaneously in all subpopulations. Even if they are governed by the same dynamical equations, they are asynchronic, which means that the onset of the outbreaks in the subpopulations are not necessarily simultaneous [20], [21], [22]. Adding the corresponding differential equations of all subpopulations one gets

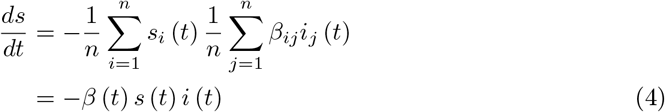

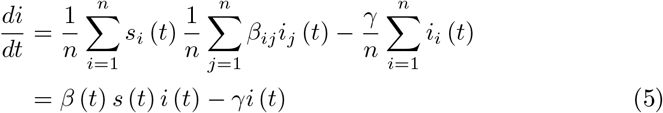

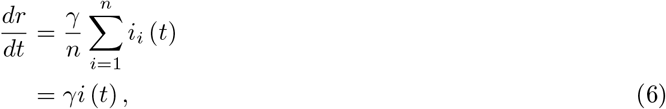

where 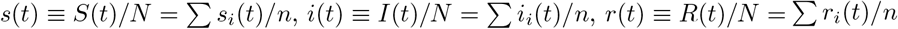,are the fractional susceptible, infectious and removed individuals of the total population with *N* individuals and *N = nN_i_* relates the total population with that of each subpopulation. To obtain the final relations (4)-(6) we have defined a time-dependent transmission rate

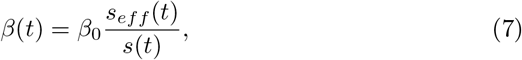

with

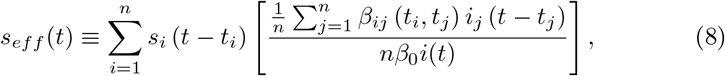

where we have considered that the difference between subpopulations is due to different onset times *t_i_*. If all the subpopulations were identical and synchronic then *β_ij_ = β*_0_ and we recover the standard SIR result *β* (*t*) *= β*_0_. However, in general, *β* (*t*) *< β*_0_ when subpopulation onset times do not coincide. Thus, asynchronicity has been incorporated in a SIR framework through a time dependent transmission rate *β* (*t*). As we will show below, a time-dependent transmission rate may be used to explain the early subexponential growth of the spread, even in the absence of susceptible depletion or intervention measures. Let us stress that other SIR models also consider time-dependent transmission rates but in those cases it is introduced externally and mainly to model the reactive behavior of the population in response to containment measures [12], [23], [24]. In contrast, in the present model *β* (*t*) is obtained as part of the solution of the dynamic equations and its time dependence appears even if the population contact dynamics is uniform and the depletion of susceptible individuals is negligible.

## A time-dependent transmission rate that describes correctly the spread of COVID-19 and leads to early algebraic growth

The calculation of *s_ef f_* (*t*) in Eq. (7) would depend on the distribution of subpopulation onset times and is affected by fluctuations originated in the exchange of individuals between them. The effective fractional susceptibility *s_eff_* (*t*) can be thought as a weighted average over subpopulations, with the quantity in square brackets in Eq. (8) playing the role of a weight factor. Assuming that we can transform the average over subpopulations into a time average and making a continuous approximation we propose to write

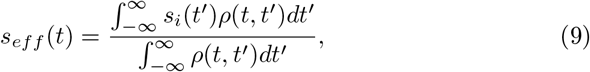

with *ρ*(*t*, *t*′) a weight function that depends on the time t, when the average is calculated, and the time *t*′ measured from the time the first case appeared in a given subpopulation. Lacking additional information about the distribution of onset times, population sizes, or mixture rules, here we make the parsimonious assumption that *ρ* (*t* − *t*′) = Θ (*t*′ *− t*′) Θ (*at*′ − *at*_0_), with Θ (*x*) the Heaviside step function. In other words, the average given by Eq. (9) has been approximated by a set of local outbreaks with a uniform distribution of onset times, between an initial time *at*_0_ and a final time *at*,

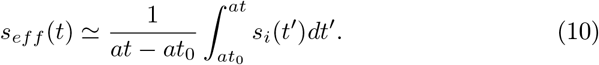

This approximation pretends to describe the effects on the transmission rate, of the individual asynchronic subpopulations that form the original inhomogeneous system by an equivalent effective system in which the number of susceptible individuals is the mean of local outbreaks with a uniform distribution of onset times. The value of the parameters *t*_0_ and *a* are obtained from the fitting to the data points of the epidemic curve for a given population. Negative values of *t*_0_ incorporate the contributions of the subpopulations that started their local outbreaks before time *t* = 0 (see Materials and Methods for the definition of the initial time) and their contribution to the average decreases as time *t* increases. The parameter *a* is a measure of the *strength* of the asynchronicity of the onset times due to a exchange of individuals between subpopulations. A small value of *a* means a diminished dispersion of the onset times of the local outbreaks and conversely, larger values of *a* means a larger dispersion of starting times. This quantity is related to the rate of propagation of the infectious agent between different subpopulations, the larger the delay of the transmission, the larger the parameter *a*. The fact that the upper limit in the integral of Eq. (10) depends on *t* means that as the time goes by, the dispersion of the local outbreaks increases. When *at → at*_0_ essentially all the local outbreaks are at the same stage and thus initially, the global outbreak has an standard SIR character and we expect an initial exponential growth.

To examine the validity of the proposed approximation, we apply it to the COVID-19 epidemics in different countries starting with the case of Mexico. Figure 1 (top panel) shows a semi logarithmic plot of *s* (*t*) and *i* (*t*), as obtained from Eqs. (4)-(5), and compare with the observed data (by confirmation date) [25] for both, the total number of infected (cumulative cases), 1 *− s* (*t*), and infectious individuals (active cases), *i* (*t*), for the case of Mexico. In practice, active cases correspond to individuals whose symptoms started within the previous 14 days from the day a given data point was released. An initial basic reproduction number *R*_0_ *=* 2.2 and recovery rate γ =1/6 were used as parameters for phase 1, that is, before containment measures were adopted. After *t =* 20 days, once containment measures were adopted and their effects started to be noticeable, the initial reproduction number was changed to *R*_0_ *=* 1.7. The data have been plotted from March 9th, which corresponds to *t =* 0, where the data points show the beginning of a regular behavior (see Materials and Methods). The bottom panel shows the empirical new daily cases and the model fit. Remarkably, the model is able to reproduce correctly the empirical infectious cases (yellow line) and the new daily cases data, considering that the fitting was performed only for the total number of infected (blue line), no independent fittings for each curve were required. Figure 2 shows the early growth of the total number of cases. After a first short exponential growth, an algebraic dependence with scaling law ≃ *t*^3.4^ follows. Let us stress that, in contrast to other approaches [14], [15], this scaling law appears exclusively due to the time dependence of the transmission rate *β* (*t*) arising from the assumption of asynchronic transmission of the local outbreaks even without reactive population behavioral changes and before susceptible depletion sets in. Figure 3 shows with lines the behavior of *β*(*t*) as obtained from the model, Eq. (7) with the approximation given by Eq. (10). The first section of this curve (blue line) corresponds to the beginning of the outbreak, when propagation was free, without containment measures. The second section of the curve (yellow line) corresponds to days after containment measures were implemented. In both cases, the transmission rate decreases with time. The data points are obtained from Eq. (4) employing real data for Mexico [25]. We used the fact that new daily cases equals *−ds/dt*. The grey vertical line signals the end of the generalized voluntary quarantine (“Jornada Nacional de Sana Distancia”) and mitigation measures started to be released locally, depending on the strength of the epidemic in each state of the country. Accordingly, rebounds are possible after this date. Other approaches [12], [24], [26], attribute the time-dependence of *β*(*t*) to the reactive behavior of the population in response to non-pharmaceutical mitigation measures and is introduced externally to the model. In contrast, in the present model, the temporal variation of *β*(*t*) is attributed to the asynchronicity of the outbreaks due to a delay in the exposure to the pathogen, of individuals in different subpopulations even if the behavior of the population remains unaltered and is obtained from the model dynamical equations. The right axis shows the corresponding values of the basic reproduction number [23], *R*_0_ (*t*) *= β* (*t*) /γ.

**Figure 1:**
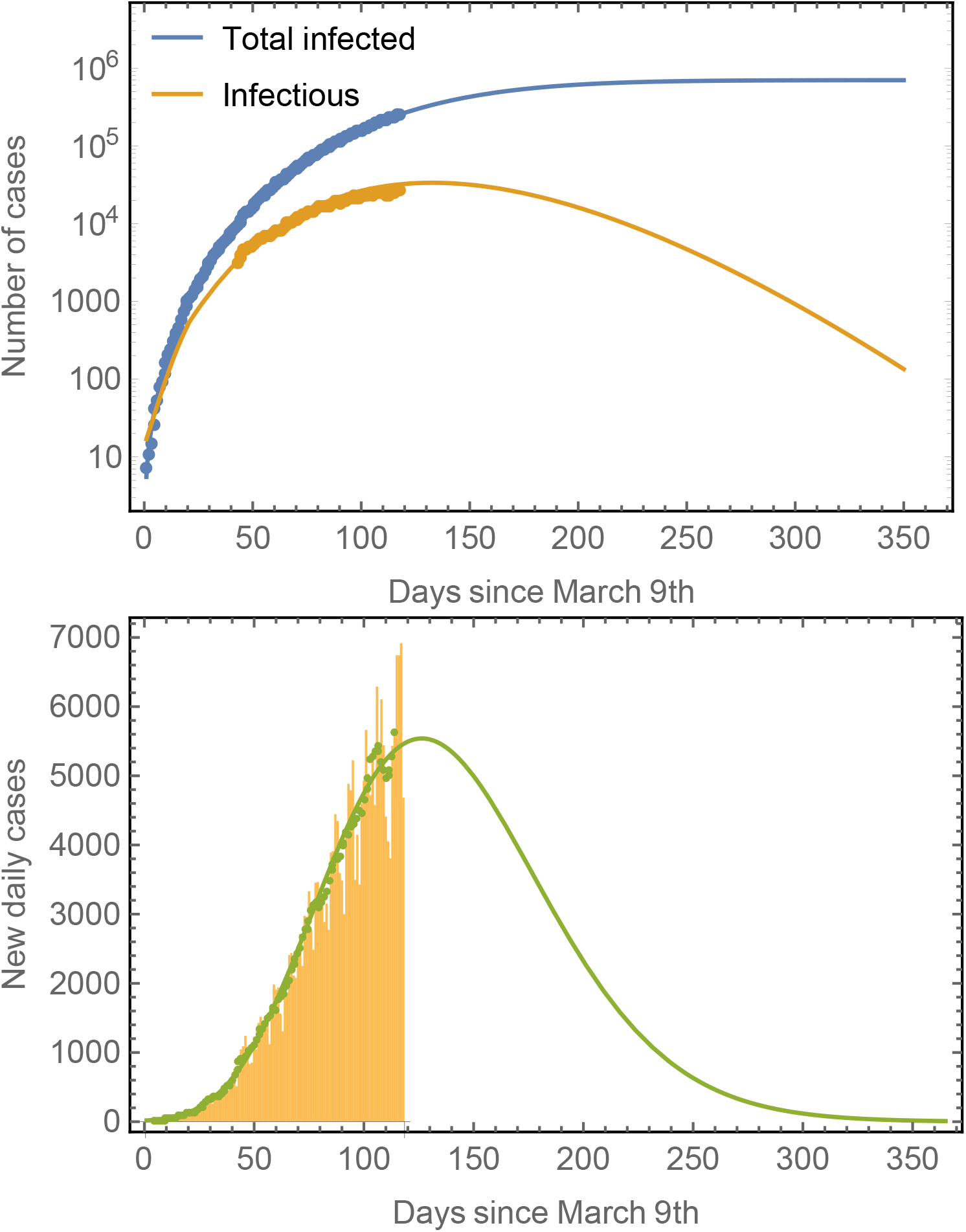
Number of cases in Mexico compared to model predictions. The total number of cases (blue line) is obtained from fits to the model defined by Eqs. (4)-(10) to real data for Mexico. The prediction of the number of infectious individuals and of the new daily cases is obtained as a consequence, no independent fittings are required. The model predicts that the peak time for the number of infectious cases is around July 24th whereas that for the new daily cases is around July 18th. The number of total infected saturates at around 800,000. Note that is a common trend in most epidemiological models that the long-time predictions tend to underestimate final observations. Predictions improve as the number of data points considered in the fitting increases. The green symbols in the right panel correspond to seven-day averages of the new daily cases data represented by the yellow bars.

**Figure 2:**
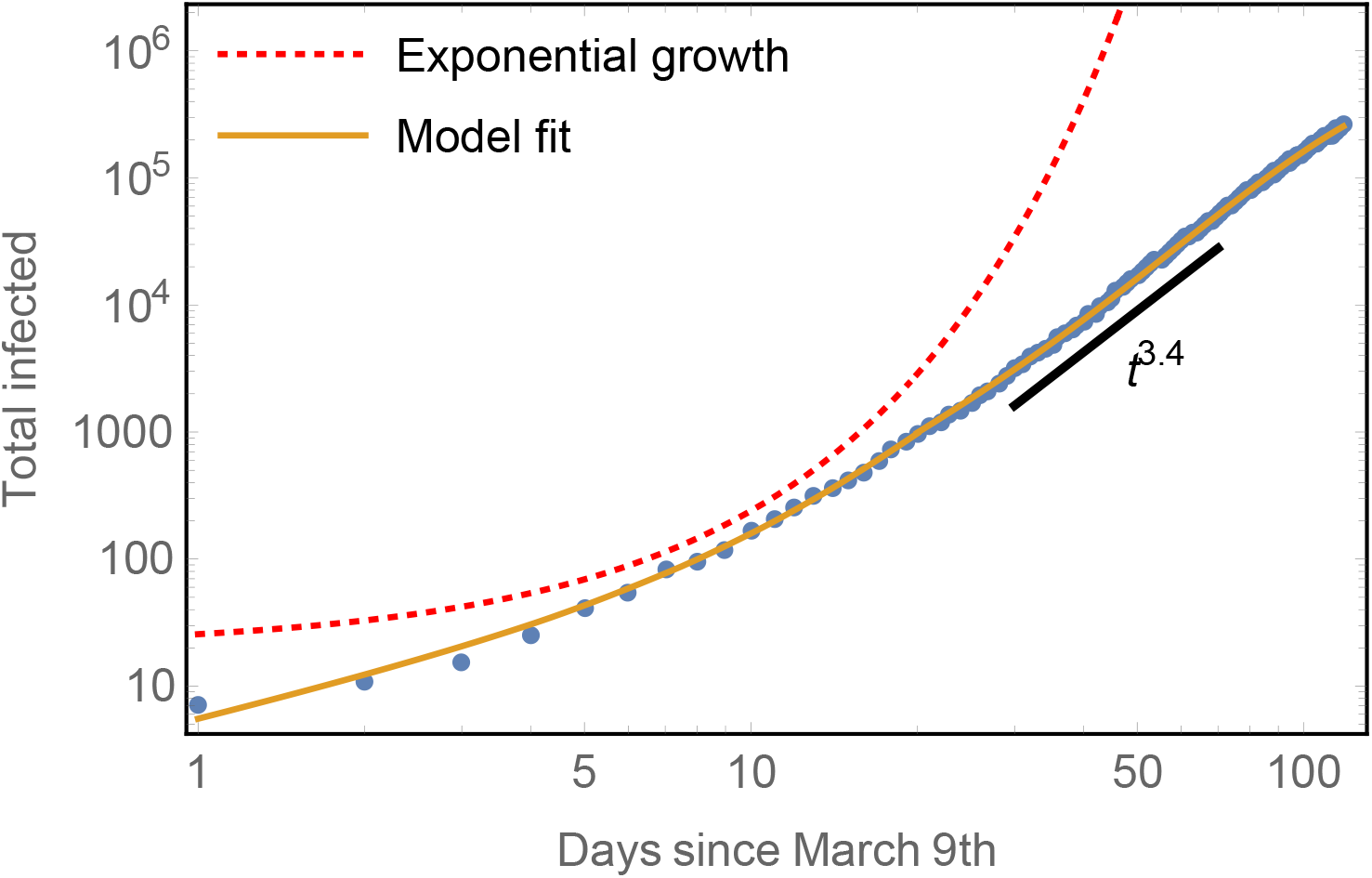
Early subexponential growth of the total number of cases compared to model predictions. The model captures well both, the initial exponential rise of total infected as well as the subsequent algebraic growth with scaling ≃ *t*^3.4^. An example of exponential growth is shown for comparison.

**Figure 3:**
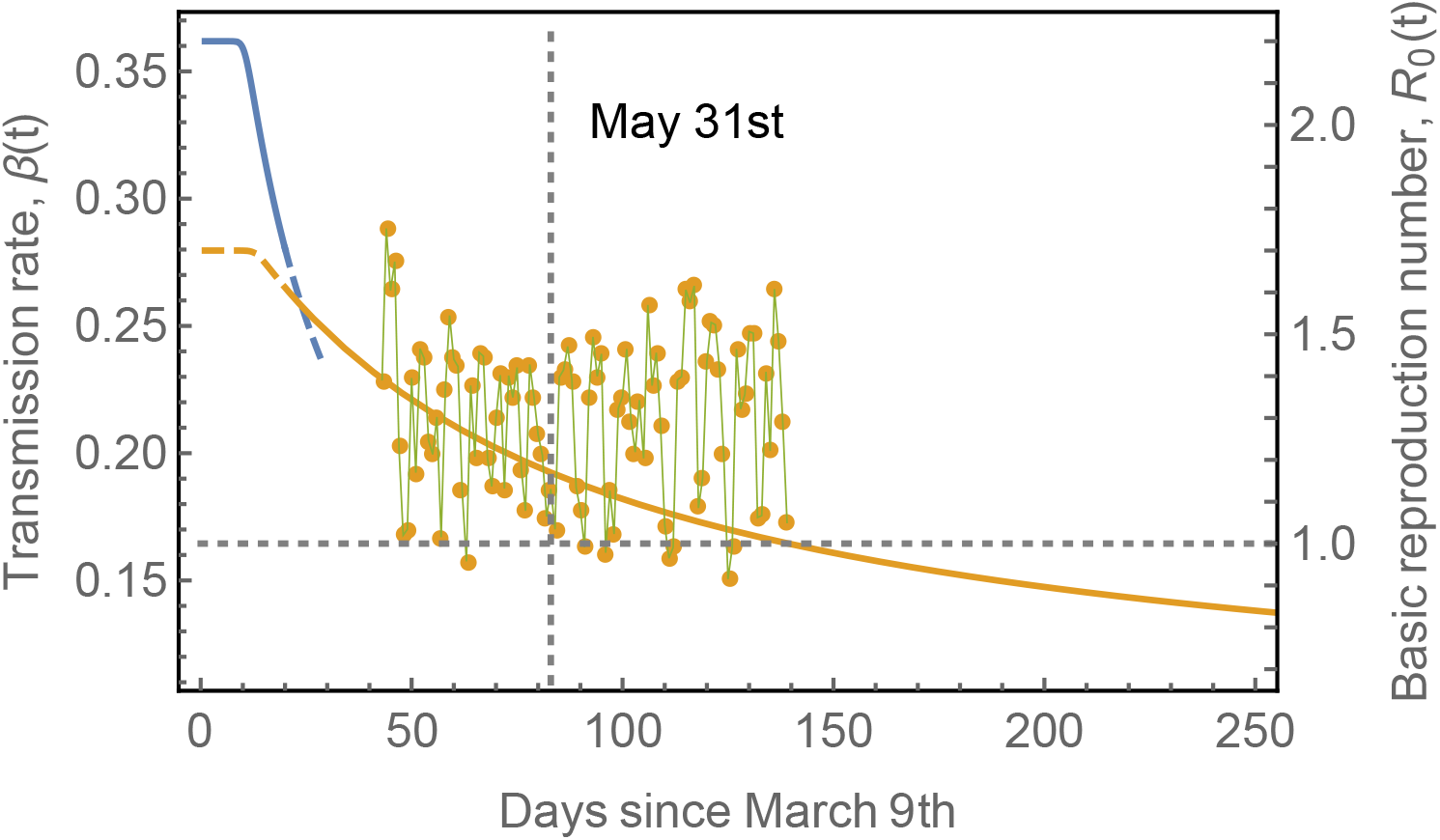
Transmission rate, *β*(*t*) and basic reproduction number *R*_0_ (*t*) = *β* (*t*) /γ. Two different sections of the transmission rate as direct consequence of containment policies. The initial blue section corresponds to a situation in which no specific policy was considered and the spread proceeds without restrictions. The yellow line is the result of the containment measures, consisting in the closure of non essential activities and the suggestion to stay at home starting from April 1st. The time dependence of *β*(*t*) is not due to the containment policies but is a natural consequence of the asynchronicity of the subpopulation spreads. However, once containment measures were adopted, the basic initial reproduction number decreased from *R*_0_ (*t* = 0) = 2.2, to around *R*_0_ (*t* = 0) ≃ 1.7. The symbols are obtained from Eq. (4) using real data for Mexico. The dashed vertical line marks the end of the generalized voluntary quarantine of the population at May 31st. The recovery rate was γ = 1/6.

Since the first outbreak of COVID-19 in Mexico remains in progress, it is interesting to evaluate the performance of the model in countries where the first outbreak has nearly ended. We have chosen the eight European countries with the largest number of COVID-19 cases to further validate the model [27]. Figure 4 shows the epidemic curves for Belgium, Italy, France, Germany, Netherlands, Spain, Sweden, and United Kingdom. The model-fits capture surprisingly well the epidemic progression in all cases in spite of the different mitigation strategies applied by each country [28], [29].

**Figure 4:**
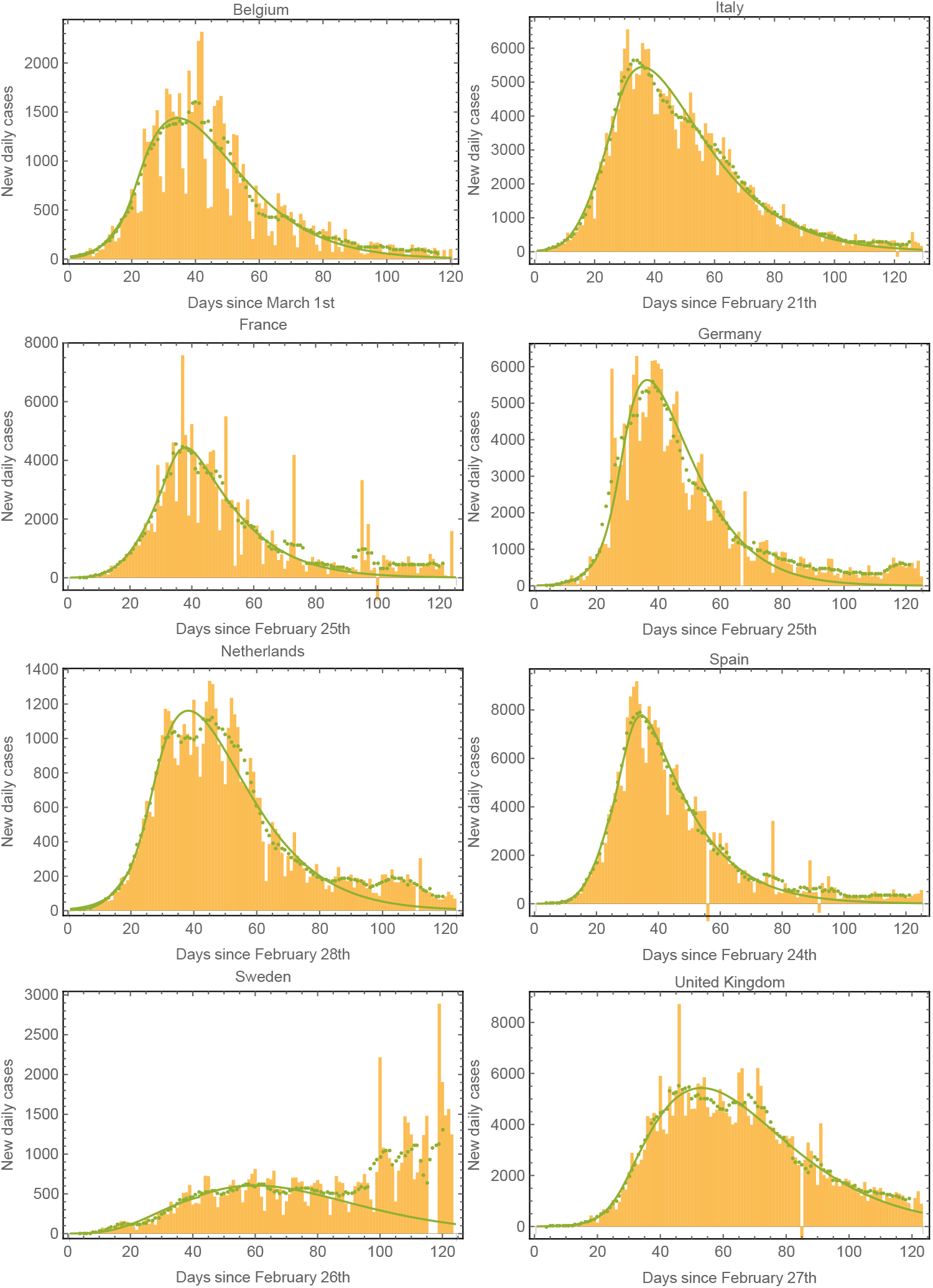
Epidemic curves for various countries. We plot the data and model fits for the new daily cases for the eight European countries with the largest number of cases. The model fits capture correctly the epidemic progression in all cases. The green symbols correspond to seven-day averages of the data represented by the yellow bars.

Table 1 shows the parameters used to fit the model in Fig. 4. In most of these countries, containment measures were taken at about 20 *−* 40 days after time *t =* 0 days, and thus the epidemic curve can be divided in two sections, one before and one after the containment measures were adopted. The positive values of *t*_0_ approximately reflects the time where mitigation measures were adopted. In contrast, the case of Mexico shows a negative value of *t*_0_ after the containment measures were adopted, reflecting the fact that the measures were taken very soon in the epidemic progression. This is also consistent with the fact that the evolution of the outbreak is taking considerably longer in Mexico than in most of the other countries analyzed.

**Table 1:** Fitting parameters used in Fig. 4. The effects of containment measures start to be observed at times *t* ≃ 20 − 40 days (labeled approx. transition day). Parameters are rough estimates and should not be considered accurate values.

|  |  | Before containment |  |  | Transition day | After containment |  |  |
| --- | --- | --- | --- | --- | --- | --- | --- | --- |
| Country | $\gamma$ (1/days) | $R_0$ | $t_0$ (days) | $a$ | | $R_0$ | $t_0$ (days) | $a$ |
| Belgium | 1/6 | 2.5 | -15.53 | 6.88 | 20 | 2.0 | 12.47 | 4.05 |
| Italy | 1/6 | 2.5 | -9.09 | 5.48 | 29 | 1.7 | 19.37 | 5.01 |
| France | 1/6 | 2.8 | -13.55 | 5.47 | 21 | 1.8 | 32.43 | 3.50 |
| Germany | 1/6 | 2.5 | -53.45 | 7.62 | 24 | 2.25 | 23.55 | 2.82 |
| Netherlands | 1/6 | 2.7 | -6.86 | 5.01 | 17 | 2.0 | 18.29 | 3.4 |
| Spain | 1/7.5 | 3.7 | -2.19 | 3.52 | 29 | 2.15 | 28.77 | 3.37 |
| Sweden | 1/6 | 2.5 | -9.16 | 5.07 | 42 | 1.8 | 6.69 | 5.22 |
| United Kingdom | 1/6 | 2.3 | 1.53 | 3.28 | 36 | 1.7 | 16.47 | 4.29 |

## Discussion and conclusion

Summarizing, it has been shown that the spread of the local outbreak onset times can be incorporated in a SIR formulation through the use of a time dependent basic reproduction number. This quantity is obtained as a solution of the dynamical equations and not introduced externally. Thus, its time dependence does not arise as a result of changes in social behavior in response to containment measures or of the depletion of susceptible individuals. We have shown that a simple assumption for the distribution of onset times on the calculation of the transmission rate can be at the origin of the algebraic growth observed at the early stages of the COVID-19 outbreak. This contradicts the common assumption that the early growth phase should be exponential in the absence of susceptible depletion or interventions measures. In the present model, containment measures contribute by decreasing the initial reproduction number or equivalently, the initial transmission rate. Other epidemic outbreaks also show early subexponential growth and a number of potential mechanisms have been proposed to explain it. Among them are the spatial heterogeneity and clustering of contacts arising from the fact that the number of non-infected individuals in the immediate neighborhood of infecting agents is strongly constrained [31], [32]. Reactive population behavior has also been proposed to explain changes that can gradually mitigate the transmission rate [14], [19], [24], [33]. Related to these mechanisms, a range of mechanistic models have been proposed that can reproduce subexponential growth dynamics before susceptible depletion sets in. These includes models with gradually declining contact rate over time [12] and spatially structured models such as household-community networks [19], among others. However, for real epidemics, the underlying mechanisms governing subexponential growth has be difficult to disentangle and the matter remains debated [19], [30], [31].

The model predicts a final number of cumulated cases that is substantially smaller than that predicted for homogenous well-mixed populations in agreement with models that predict smaller disease-induced herd immunity when population heterogeneity is taken into account [16].

The present model was validated using the case of the COVID-19 outbreak in different countries with an emphasis in the case of Mexico. As of the origin of the asynchronicity one can propose different underlying mechanisms, including the transmission between geographical dispersal subpopulations as individuals travel among them [5] [7], [13]. Also, the existence of social structures that allow to divide the community in different social cohorts with transmission between them due to contact patterns that are lower than that between individuals of the same cohort [16], [34]. In general, the overall connectivity between subpopulations will determine to a large extent the rate of propagation of the viral agent and the final number of cases. Subpopulations more efficiently connected between them will have smaller dispersion of onset times. The effect of the strength of the asynchronicity is shown in Fig. 5 which shows the evolution of the total number of infected, considering different values for the parameter *a*, for a given *R*_0_. We observe that the long-time total number of infected individuals will be highly dependent on *a*. The blue line corresponds to the fitted value of *a* for the case of Mexico while the yellow and green lines are the predictions for a smaller (smaller *a*) or a larger (larger *a*) asynchronicity.

**Figure 5:**
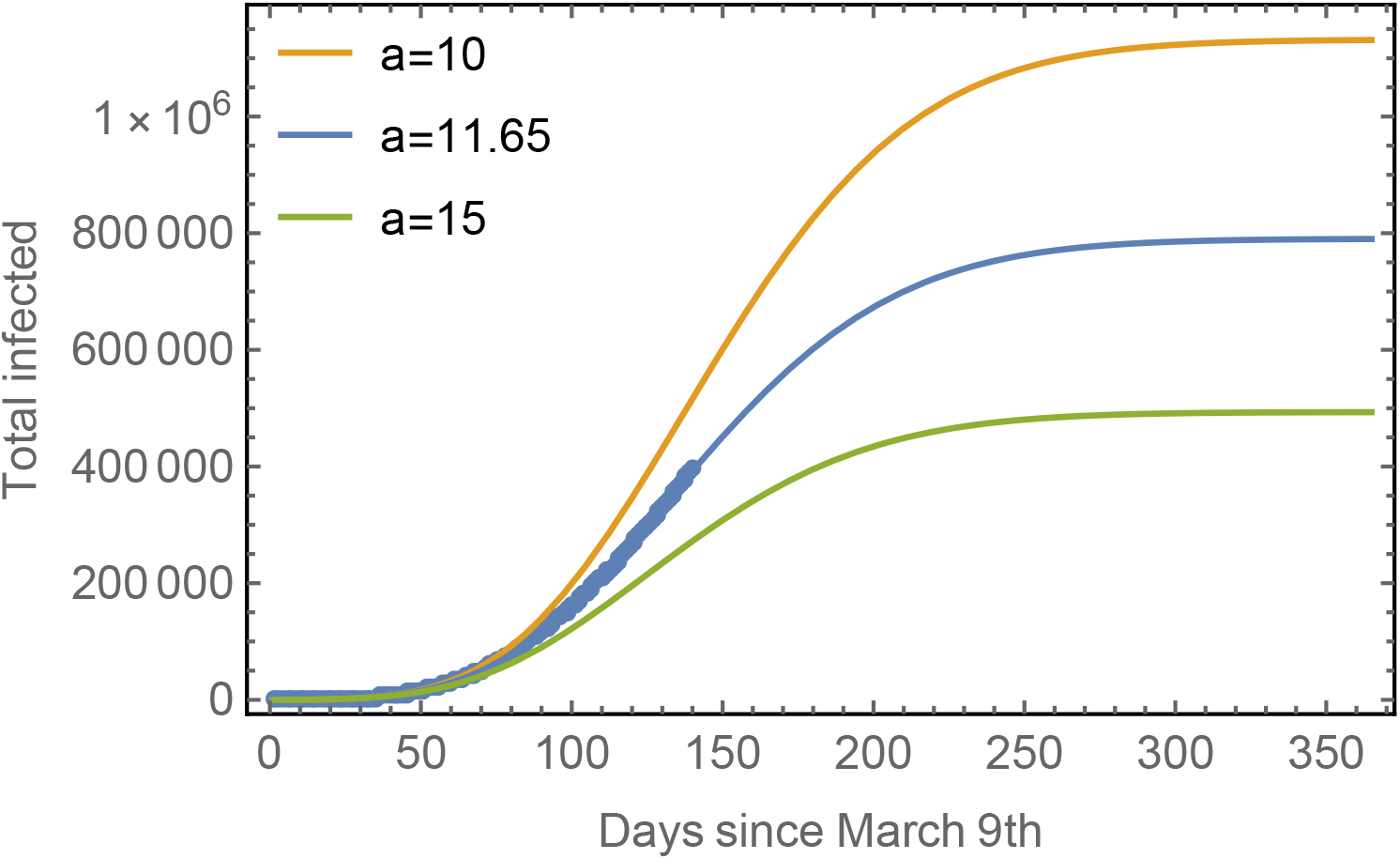
Progression of the spread assuming different asynchronicities. Blue line: The observed data points for the total number of cases in Mexico with the fitted model. Yellow line: The predicted evolution for subpopulations with a smaller asynchronicity (*a* = 10). Green line: The predicted evolution for subpopulations with a larger asynchronicity (*a* = 15). In all cases, *R*_0_ = 1.7.

By reducing the number of infectious individuals traveling between subpopulations, the probability of a susceptible individual to get infected is reduced. Analogously, an infectious individual that takes more time to travel from one subpopulation to another (increasing asynchronicity), will have more time to recover before infecting a susceptible individual in the destination subpopulation. In this sense, this work is consistent with mitigation strategies consisting in the design of containment mechanisms oriented to increase asynchronicity (or, relatedly, reducing connectivity), for example, imposing travel restrictions for long-distance routes, partially isolating subpopulations from each other or by increasing the time an individual takes to move from one subpopulation to the other. Furthermore, travel restrictions could be targeted to highly-connected individuals that travel to several subpopulations. For countries where an strict quarantine is impracticable, this could be a more realistic alternative. We believe that the insight obtained from the present model may be useful for planning non-pharmaceutical responses to better mitigate or block the overall spread of an epidemic outbreak.

## Materials and Methods

Identifying the first infectious case is a difficult task so that considering *t* = 0 days as the day the first case was reported does not seem to be appropriate. For this reason, before trying to fit the original data points, we first shifted them in time so that the time *t* = 0 days corresponds to the point where a discernible regular behavior starts to appear. This is shown in Fig. 6 where the data for Mexico that were disregarded for fitting purposes correspond to dates ranging from February 27th, the day of the first detected case, until March 9th. Those data show an irregular behavior probably due to the few number of cases and large relative fluctuations.

**Figure 6:**
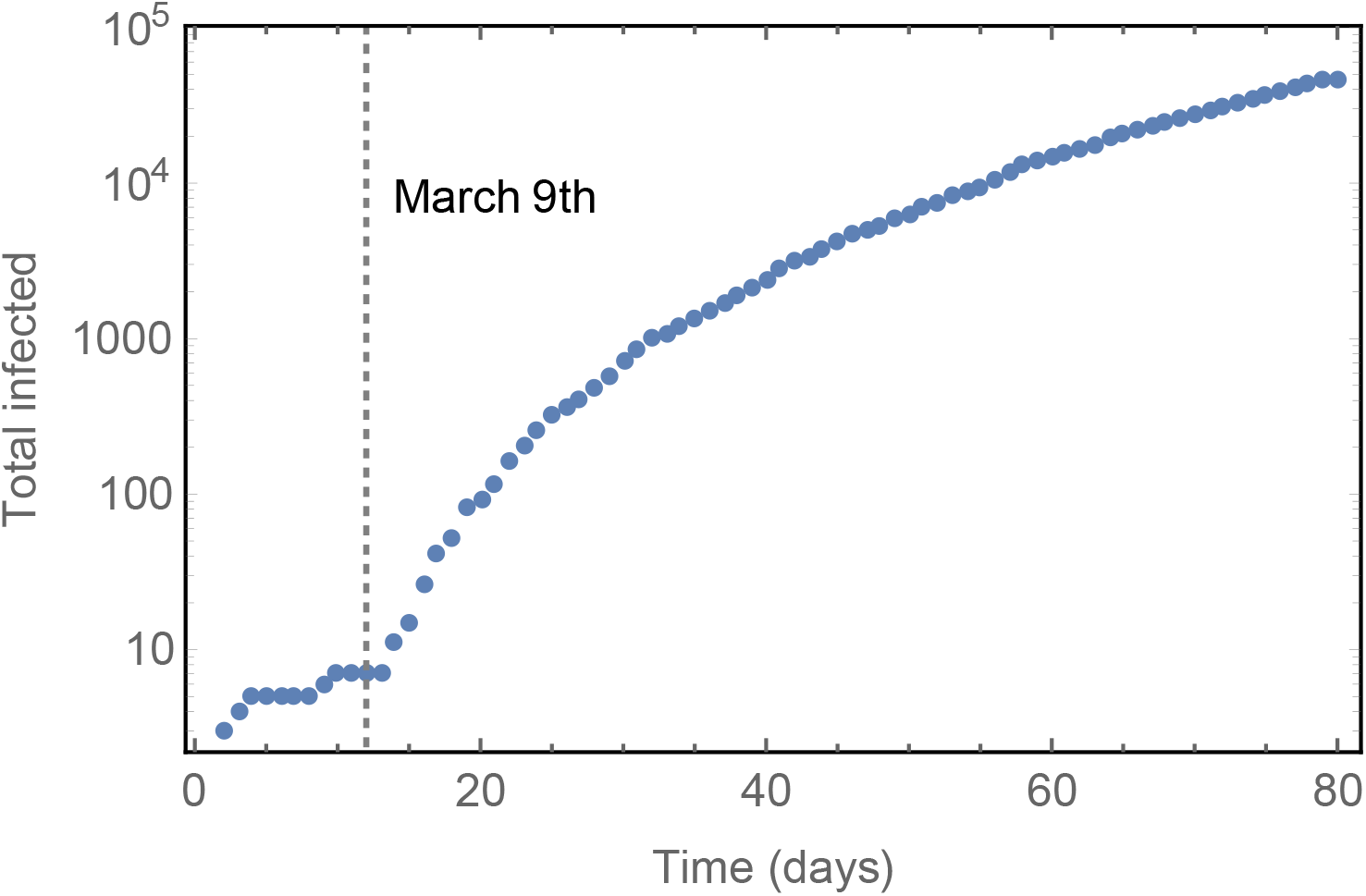
Time shift of the data for Mexico. The data for the total number of infected shifted in time so that at *t =* 1 day, the first discernible data corresponding to a regular behavior. In this case the first regular point appears on March 9th. Then, the first detected case (the “zero patient” of the total population) appears at time *t = −*12 days, that is, on February 27th.

The exact solution for *s_i_* (*t*) used in Eq. (10) should be obtained from Eqs. (1)-(2) but the quantities *β_ij_* are unknown. Here we approximate *β_ij_* ≃ *β*_0_*δ_ij_*, with *δ_ij_* the Kronecker delta function such that *s_i_* (*t*) would be the solution of a standard SIR model. Then, the obtained *s_i_* (*t*) could be directly substituted into Eq. (10). From a practical point of view, it is numerically easier to approximate *s_i_* (*t*) by the function

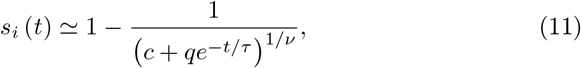

where the parameters *c, q*, *ν*, and *τ* are obtained from the fitting to the exact form for *s_i_* (*t*) once *β*_0_ and γ are known. The common initial conditions *s_i_*(0) = 1 − 1/*N_i_*, *i_i_*(0) = 1/*N_i_*, and *r_i_*(0) = 0 were assumed. Figure 7 shows an example for *s_i_* (*t*) as obtained from standard SIR model (solid line) and its approximation Eq. (11) (dashed line). By incorporating Eq. (11) into Eq. (10) and this last result into Eqs. (4) to (7), the evolution of *s* (*t*)*, i* (*t*), and *r* (*t*) for the whole population is finally obtained.

**Figure 7:**
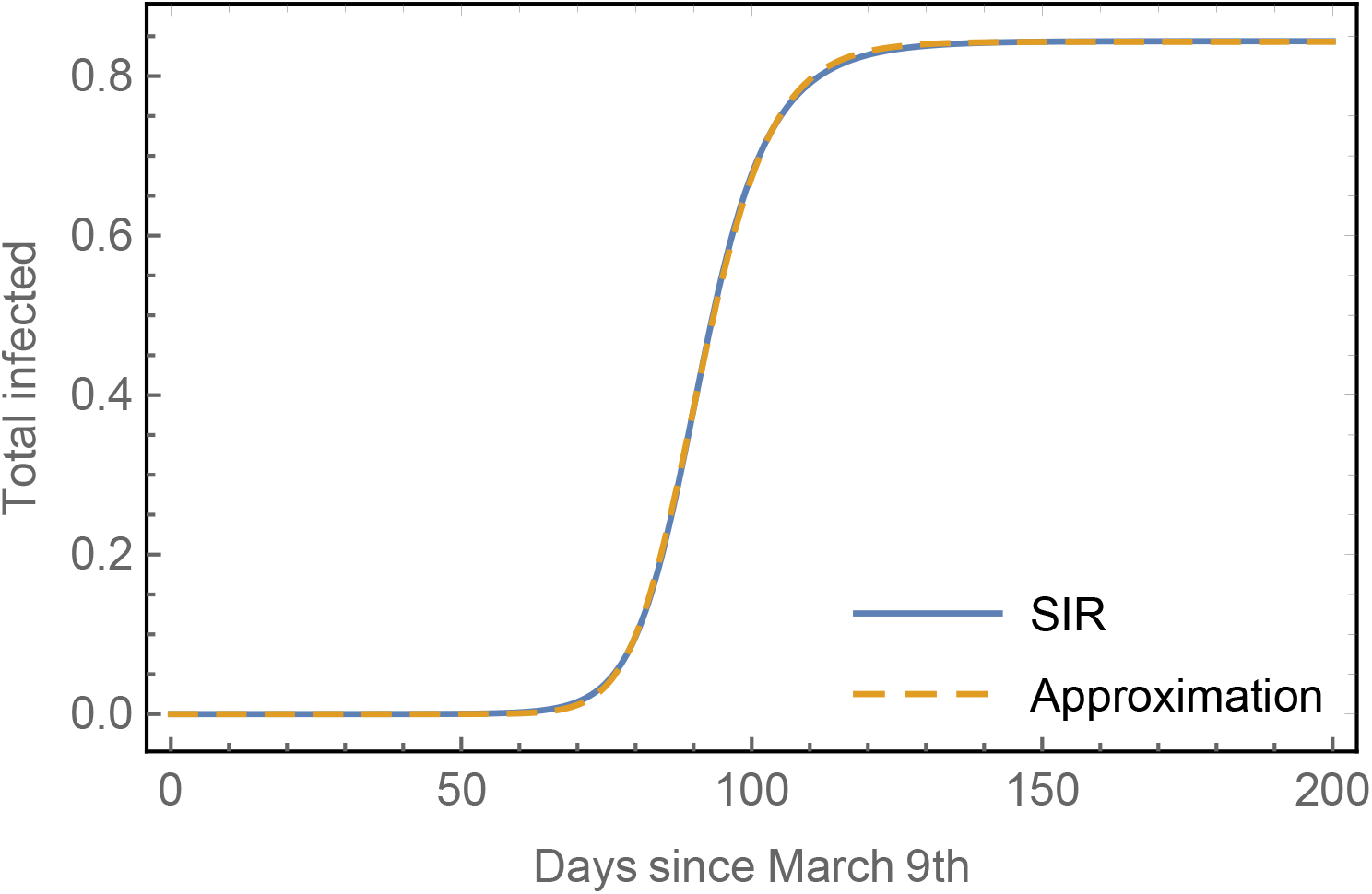
Total number of cases for a given subpopulation. The fraction of total infected (1 − *s_i_*(*t*)) is obtained from the standard SIR model, Eqs. (1) to (3), considering one infectious individual at *t* = 0 (the “zero patient” for that subpopulation) and the rest of the subpopulation considered as susceptible. The dashed yellow line is the approximation given by Eq. (11)

The fitting of the model to the data, before containment measures were taken, was done choosing as initial conditions the value of a spline function that softens the data taken arbitrarily at time *t =* 4 days, as shown in Fig. 8. In this way, possible misfitting due to fluctuations of the data at the very first data points is minimized. The fitting of the second section of the model, that is, once containment measures were taken at day 20 in the case of Mexico, considered as initial values the model values of the first section taken at day 20.

**Figure 8:**
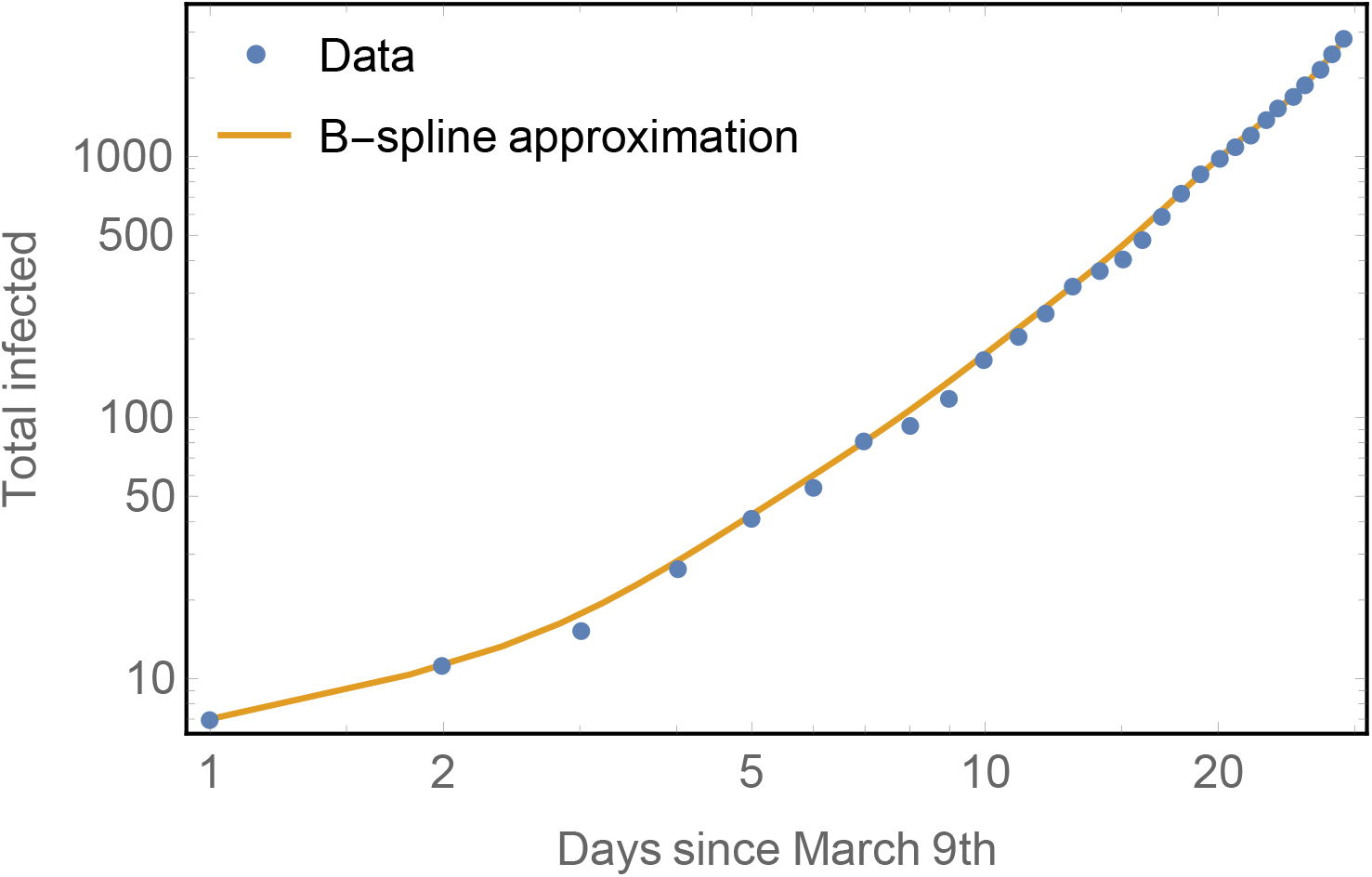
Smoothing the curve. The observed data points for the total number of cases of phase 1 are approximated using a B-spline approximation with underlying polynomial basis degree (*d*) equal the number of data points −1. In this case we used 29 data points which means *d =* 28. The value of the curve at *t =* 4 days was arbitrarily chosen as initial condition.

For the case of Mexico, the parameters used were *R*_0_ *=* 2.2 and γ = 1/6, values generally accepted for the initial regime without containment measures and *R*_0_ *=* 1.7 for the second regime after containment measures were adopted. The first fitting process considered the first data points up to *t =* 20 days with obtained fitting parameters *t*_0_ ≃ *−*15 days and *a* ≃ 8.7. Analogously, the fitting for the second regime consisting in taking data from *t =* 21 days to the last data point available. For this second regime the obtained fitting values were *t*_0_ ≃ *−*64 days and *a* ≃ 10. The model is robust to minor changes in other parameters used for the fitting, in particular, the results were largely insensitive to variations in the exact day of transition between the two regimes.

## Data Availability

All data are available under request.

## Acknowledgements

The author thanks Oleg Kogan, David Reguera and Juan Adrián Reyes for fruitful discussions and helpful comments. Partial financial support provided by DGAPA-UNAM through grant DGAPA IN-103419.

